# Gestational diabetes as a risk factor for GBS maternal rectovaginal colonization: a systematic review and meta-analysis

**DOI:** 10.1101/2023.11.02.23297989

**Authors:** Vicki Mercado-Evans, Jacob J. Zulk, Zainab A. Hameed, Kathryn A. Patras

## Abstract

**Background:** Maternal rectovaginal colonization by group B *Streptococcus* (GBS) increases the risk of perinatal GBS disease that can lead to death or long-term neurological impairment. Factors that increase the risk of rectovaginal GBS carriage are incompletely understood resulting in missed opportunities for detecting GBS in risk-based clinical approaches. There is a lacking consensus on whether gestational diabetes mellitus (GDM) is a risk factor for rectovaginal GBS. This systematic review and meta-analysis aims to address current conflicting findings and determine whether GDM should be clinically considered as a risk factor for maternal GBS colonization.

**Methods:** Peer-reviewed studies that provided GDM prevalence and documented GBS vaginal and/or rectal colonization in women with and without GDM were included in this analysis.

From study inception to October 30, 2023, we identified 6,275 relevant studies from EMBASE and PUBMED of which 19 were eligible for inclusion. Eligible studies were analyzed and thoroughly assessed for risk of bias with a modified Newcastle-Ottawa Scale that interrogated representativeness and comparability of cohorts, quality of reporting for GDM and GBS status, and potential bias from other metabolic diseases. Results were synthesized using STATA 18 and analyzed using random-effects meta-analyses.

**Results:** Studies encompassed 266,706 women from 10 different countries, with study periods spanning from 1981 to 2020. Meta-analysis revealed that gestational diabetes is associated with a 16% increased risk of rectovaginal GBS carriage (OR 1.16, CI 1.07-1.26, *P* = 0.003). We also performed subgroup analyses to assess independent effects of pregestational vs. gestational diabetes on risk of maternal GBS carriage. Pregestational diabetes (Type 1 or Type 2 diabetes mellitus) was also associated with an increased risk of 76% (pooled OR 1.76, CI 1.27-2.45, *P* = 0.0008).

**Conclusions:** This study achieved a consensus among previously discrepant observations and demonstrated that gestational diabetes and pregestational diabetes are significant risk factors for maternal rectovaginal carriage of GBS. Recognition of GDM as a risk factor during clinical decisions about GBS screening and intrapartum antibiotic prophylaxis may decrease the global burden of GBS on maternal-perinatal health.

## INTRODUCTION

Group B *Streptococcus* (GBS) remains a leading cause of neonatal morbidity and mortality across the globe despite nearly two decades of systematic implementation of preventative approaches that include universal maternal screening or risk-based administration of antibiotics during delivery[1]. GBS colonizes the vaginal and/or gastrointestinal tract of about 18% of pregnant women[2, 3].Neonates can acquire GBS during passage through the vaginal canal during delivery, and GBS also has the capacity to ascend into the uterine-fetal space before parturition. While some women and their neonates are colonized without symptoms, GBS can cause devastating complications and disease such as spontaneous abortion, preterm labor, stillbirth, and neonatal sepsis and meningitis[1, 4]. The mechanisms driving the divergence in GBS pathogenic vs. commensal behavior is poorly understood. Currently, the following maternal factors are clinically recognized for increasing the risk of GBS neonatal disease and are used to identify women who should be given intrapartum antibiotic prophylaxis (IAP): 1) previous infant with early-onset GBS disease (EOGBS), 2) GBS bacteriuria during the current pregnancy, 3) temperature > 38 °C during labor, 4) prolonged rupture of membranes (PROM) > 18 hours, or 5) delivery at < 37 weeks of gestation[5]. Some countries practice this risk-based approach and others implement universal screening of women around 35 weeks of gestation, with subsequent IAP for those who have rectovaginal GBS carriage. Screening based approaches are associated with enhanced protection against neonatal GBS early onset disease (EOD; occurring in the first week of life) compared to risk-based strategies[5], which suggests that we have yet to understand all of the maternal factors that predict GBS disease. Additionally, up to 46% of cases of EOD occur in the absence of the risk factors currently used for clinical decision making[5–7].

One possible additional risk factor for maternal rectovaginal GBS colonization is gestational diabetes mellitus (GDM) which affects approximately 14% of pregnancies worldwide[8]. GDM, diabetes that acutely develops during pregnancy, is a state of heightened insulin resistance, insufficient pancreatic insulin production, hyperglycemia, immune dysregulation, and altered vaginal microbial composition[9–12]. This systemic disruption to maternal physiology leads to an increased risk of complications including preterm birth, pre-eclampsia, and a long-term increased risk of cardiovascular disease in both women and their children. In clinical cohort studies, infants born to women with gestational diabetes are at greater risk of early onset culture-verified GBS sepsis[13], late onset clinical sepsis[14] and extended hospital stay[15]. Considering that rectovaginal GBS carriage is the primary risk factor for GBS neonatal sepsis, women with GDM may have greater GBS colonization rates thereby imparting increased risk of neonatal disease. Observational clinical studies have reported conflicting findings on the association between diabetes (pregestational Type 1 or Type 2 and GDM) and GBS carriage; some have found increased risk of GBS colonization[16–21] in diabetic pregnant women (pregestational and/or gestational), while others found no association[15, 22, 23]. Several of these studies did not specifically distinguish pregestational (Type 1 or Type 2 DM) from GDM. Although these metabolic diseases share several features, the acuity and specificity to pregnancy of GDM lends unique insight into the pathogenic potential of group B *Streptococcus*.

The aim of this study was to conduct a systematic review and meta-analysis of the risk for rectovaginal GBS carriage in women affected by GDM. Resolving whether GDM is an independent risk factor for maternal GBS colonization is essential for closing the gap in current IAP approaches; a critical step towards reducing the global burden of GBS-associated neonatal morbidity and mortality.

## MATERIALS AND METHODS

### Search strategy and study selection

Studies were identified through a database search that included PubMed and EMBASE, which encompassed MEDLINE and preprints as sources (**Fig. 1**). The search strategy implemented search terms intended to capture two kinds of studies: 1) Those that specifically assessed GBS maternal colonization and/or neonatal transmission in women with gestational diabetes, and 2) studies on GBS maternal colonization prevalence which included information about gestational diabetic status and respective GBS status.

**Figure 1:**
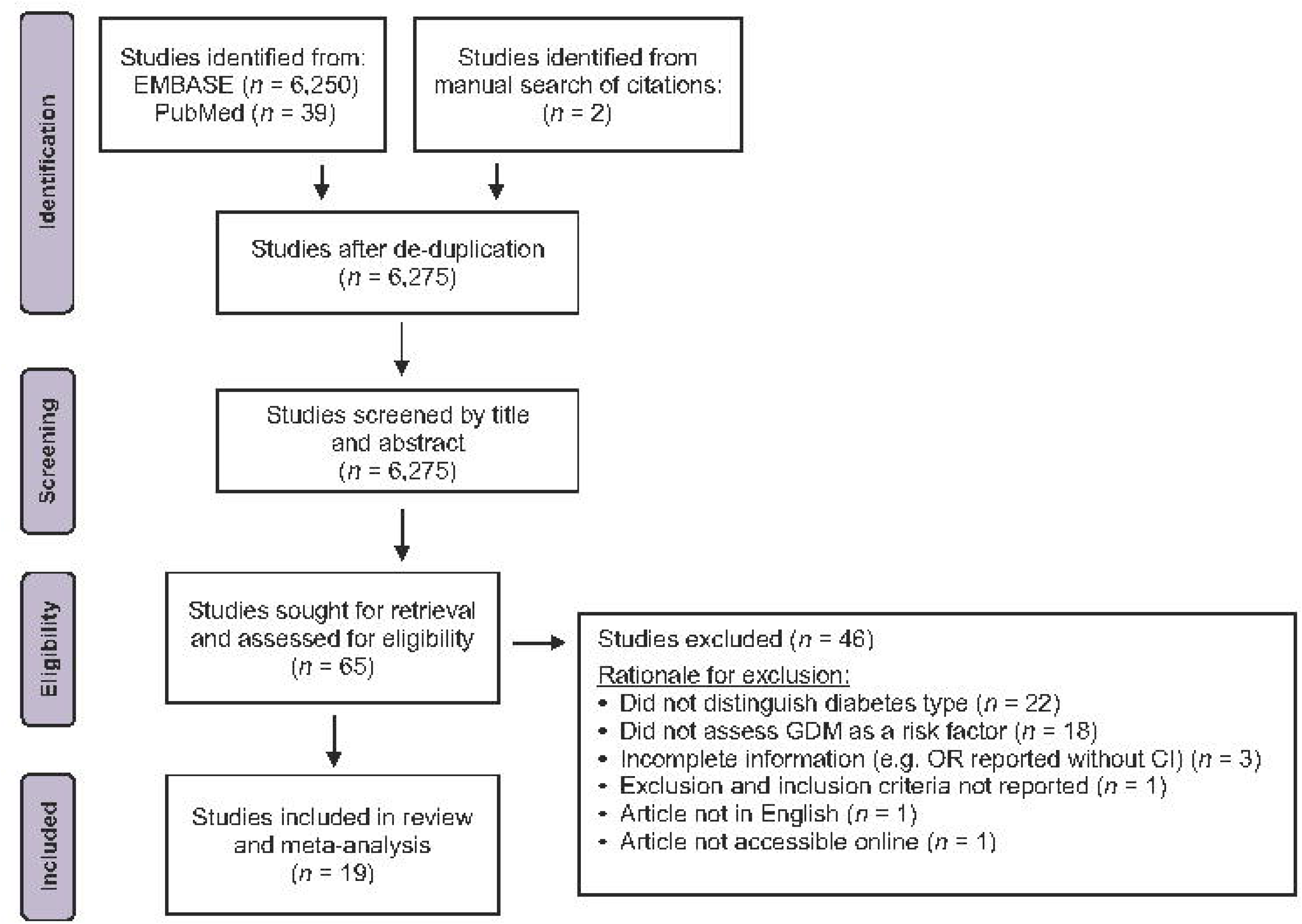
Study identification, screening, and selection process. Flow diagram of selection of the included studies.

As such, a combination of the following search terms was implemented: gestational diabetes or GDM, chorioamnionitis, newborn and sepsis or cocci and sterile site, and group b streptococcus or GBS or streptococcus agalactiae, English, humans. The following search terms were used as filters followed by the word not: in vitro, ex vivo, animal, tilapia, zebra fish, bovine, breast milk, phylogeny, case study, cells, case report, urinary tract infection, non-pregnancy, endocarditis, murine, mouse, primate. Reviews, conference abstracts and editorials were excluded from our search. The literature search was restricted to human studies published in English, with no study period restrictions. The last query was performed on 10/30/2023. Titles and abstracts were screened for adherence to the inclusion and exclusion criteria detailed below. This study was not registered, but the Preferred Reporting Items for Systematic Reviews and Meta-Analyses (PRISMA) was used as a guideline for this systematic review and meta-analysis.

### Inclusion and exclusion criteria

The inclusion criteria for this study were: peer-reviewed studies that documented GBS vaginal and/or rectal colonization in expectant mothers, with information about the proportion of women who were clinically diagnosed with GDM. Studies were required to provide the proportion of women who did not have GDM. We included studies that potentially had women with pregestational diabetes in the control, non-GDM, group but we stratified analyses accordingly. Reporting of GDM diagnosis and maternal GBS vaginal and/or rectal colonization was accepted through medical records or diagnosis from medical professionals. Studies were included irrespective of sample type used to determine GBS colonization (vaginal, rectal, or perianal region) and studies employing molecular or biochemical detection of GBS were included. Studies with self-collected vaginal swab samples (*n* = 1) were included. Additionally, observational, baseline data from interventional, case-control, retrospective and prospective, cross sectional and cohort studies were all included. The exclusion criteria for this study were: case studies, reviews, or letters to the editor, lack of a GBS negative or non-GDM control group, published in a language other than English, studies that did not explicitly state diabetes type (GDM vs. pregestational diabetes: Type 1 or Type 2 DM), or missing critical information such as exclusion/inclusion criteria or GBS and/or GDM prevalence in the study population. For relatively contemporary studies (published in the past decade) that did not specify diabetes type, we emailed corresponding authors to acquire information about the number of participants that had GDM vs. Type 1 vs. Type 2 diabetes and their GBS status. One reviewer screened titles and abstracts, and three independent reviewers screened full texts to assess studies for eligibility. Reasons for exclusion of each eligible study are provided (**Supplemental table 1**).

### Data extraction and risk of bias assessment

Two independent reviewers extracted data including: study characteristics, the number of participants in each group, and the number of GBS positive and negative cases in GDM, non-GDM and pregestational diabetic populations, odds ratios (OR), GBS detection method, GDM diagnostic criteria, gestational age at time of GBS screen and any relevant findings on maternal and/or neonatal outcomes.

All eligible studies were assessed for quality and risk of bias by two independent reviewers via an adapted Newcastle-Ottawa Scale[24] (**Supplemental table 2**) which focused on four broad criteria: 1) How representative the groups were of the greater communities from which the study was conducted, 2) comparability of the groups to each other with respect to various characteristics (maternal age, BMI, racial/ethnic representation, socioeconomic status, etc.), 3) Quality of outcome assessment (ascertainment of GBS and GDM status), and 4) Potential of bias from other metabolic diseases in each group such as obesity and pregestational diabetes (in non-GDM control group). Each assessment category was scored, and the sum was used to determine overall quality and risk of bias for each study. Any discrepancies greater than 2 points for any category were resolved via a discussion to achieve consensus. A total score ≤ 3 was considered low bias,> 3 and ≤ 6 indicated moderate bias, and ≥ 6 was classified as high risk of bias.

### Data synthesis and analysis

Unadjusted ORs were calculated for studies in which only prevalence data were provided, otherwise reported ORs for GDM or pregestational diabetes were utilized. OR calculation and analysis were performed using STATA 18. To calculate ORs, we compared the odds of rectovaginal GBS carriage in women with GDM versus women without GDM. For the sub-analysis of women with pregestational diabetes, we compared the odds of rectovaginal GBS carriage to nondiabetic women, or to women with GDM. For sub-analysis of pregestational diabetes, Piper et al., 1999[15] could not be included because this study accounted for effects of pregestational diabetes by excluding this population entirely from their study. Forest plots display prevalence, individual ORs and 95% confidence intervals (CI) and meta-analysis of pooled ORs with random effects modeling. The I^2^ index was used to assess the impact of study heterogeneity on study estimate variance[24], with low, moderate and high heterogeneity indicated by I^2^ of 25%, 50% and 75% respectively[25]. Publication bias was assessed with funnel plots of the ORs (natural logarithm) against the inverse of the standard error and Egger’s regression test, with *P* < 0.05 indicating a significantly asymmetric funnel plot and thus significant publication bias.

## RESULTS

### Study characteristics

The initial search identified 6,275 studies, of which 65 articles passed screening and were subjected to full-text assessment for eligibility (**Fig. 1**). 19 articles with study periods spanning from 1981 to 2020 were eligible based on the aforementioned criteria, with exclusion of 46 studies for various reasons (**Supplemental table 1**). **Table 1** provides a summary of study characteristics. The total number of women included in this systematic review and meta-analysis is 266,706; there were 18,715 women with GDM, 2,598 with pregestational diabetes, and 195,545 without GDM. The studied populations are representative of many communities across the globe with inclusion of Australia, Brazil, China, Finland, Lebanon, Mexico, Morocco, Nigeria, Spain, and the United States. Of these countries, none were of low-income, three were of lower-middle income, three were of upper-middle income, and four were of high-income as determined by the 2023 World Bank guidelines[26]. 6/19 were multicenter studies and routine screening for GBS colonization and administration guidelines for intrapartum antibiotic prophylaxis (IAP) was not an established practice at the time for 64% of studies. Studies also consisted of a mix of prospective (57%), retrospective (14%), cross-sectional (5%), case-control (14%), and population-based cohort (10%) study designs. Rectovaginal, vaginal and/or perineal GBS carriage was determined by culture for all studies: 14 studies performed rectovaginal sampling, 3 solely assessed vaginal carriage, and 2 studies did not specify. 5 studies performed culturing and molecular or biochemical identification as recommended by CDC guidelines whereas 14 studies had methods incongruent with guidelines or did not provide enough detail. 6 were found to have low risk of bias, 8 had moderate risk of bias, and 5 had high risk of bias (**Table 1 and Supplemental table 3**).

**Table 1:** Characteristics of studies included in this systematic review of the association between gestational diabetes and rectovaginal GBS colonization.

| Author, year | Study period | Country | Study Design | Inclusion (I) & Exclusion (E) Criteria | Mean age (SD) | GDM diagnostic criteria and ascertainment | GBS detection method | Potential confounders | Risk of bias |
| --- | --- | --- | --- | --- | --- | --- | --- | --- | --- |
| Matorras, 1988[17] | 1981-1985 | Spain | Prospective | Women randomly selected. No criteria specified. | 28.0 (6.1) | Oral GTT around 20 w. Coustan & Lewis Criteria. Pregestational diabetes based on history of diabetes or fasting glycemia >140mg/100mL. | Rectal and vaginal swabs followed by culture detection. | Of the 1,050 patients, 729 had complications that were not specified. | High |
| Raimer, 1997[27] | 11 months but year is not reported | USA | Case-control | I: All pregnant women presenting to the clinic.<br>E: HIV+, history of substance abuse, current STD. | Diabetic 30.6 (6.2)<br>non-diabetic 28.5 (6.8)<br>(*P* = 0.02) | Oral GTT 24-28 w, considered normal below 140mg/dL, abnormal screen (50 g challenge) was followed by 3h GTT (100g), two elevated values considered abnormal. | Vaginal swab followed by culture detection. | Significant difference in maternal age between diabetic and control groups. | Moderate |
| Ramos, 1997[18] | January 1995-March 1996 | USA | Prospective | I: Singleton gestation, intact membranes at enrollment, otherwise uncomplicated pregnancy.<br>E: HIV+, chronic | Diabetic 27.0 (6.5)<br>non-diabetic 24.6+/- (6.4) | Oral GTT 24-28 w. At least 2 abnormal readings: fasting glucose greater than or equal to 105 mg/dL, 1h | Rectal and vaginal swabs followed by culture detection. | N/A. Regression analyses controlled for maternal age, race, and obesity. | Low |
| | | | | steroid therapy, cervical incompetence, multifetal gestation. | ($P = 0.02$) | glucose (50g challenge) greater than or equal to 190 mg/dL, 2h glucose greater than or equal to 165 mg/dL, 3h glucose greater than or equal to 145 mg/dL. | | | |
| Piper et al., 1999[15] | January 1992-June 1994 (diabetic cohort); April 1992-December 1992 (nondiabetic cohort) | USA | Prospective | E: Women with previously affected infants and/or pregestational diabetes. | GDM 28.5 (6.2)<br>Non-diabetic 23.6 (5.5)<br>($P < 0.05$) | Abnormal glucose tolerance test with universal screening. | Rectovaginal swab followed by culture detection. | Diabetic cohort was significantly older, of higher parity, and less likely to deliver vaginally compared to nondiabetic controls. | High |
| Stapleton et al., 2005[28] | 1997-2002 | USA | Case-control | I: All singleton gestation births in Washington State.<br>E: Patients with missing data. | Cases 27.6 (6.1)<br>Controls 27.4 (6.2) | Data extracted from hospital records. | ICD codes for confirmed GBS maternal colonization or suspected carrier. | As acknowledged by authors, risk of disease misclassification and cannot distinguish women who were truly GBS negative vs. those who were not screened. | Low |
| Medugu et al., 2017[14] | May-September 2014 | Nigeria | Prospective | I: Third trimester.<br>E: Multifetal gestation, placenta previa, or elective caesarean section. | 29.8 (5) | Interviews, questionnaires, and hospital records. | Vaginal and rectal swabs and culture detection with confirmation via PathoDextra Strep Grouping kit. | Prevalence and effects of comorbidities were not considered. | Moderate |
| Chen et al., 2018[29] | January-June 2017 | Western China | Prospective | I: >35 w gestation routine prenatal care or at the time of delivery.<br>E: Multifetal gestation, GBS culture results not available. | Not reported. | Data extracted from hospital records. | Vaginal and rectal swabs and culture and biochemical detection. | Prevalence of pregestational diabetes not reported, nor other indicators of metabolic stress (BMI). | Low |
| Moraleda et al., 2018[30] | March-July 2013 | Morocco | Prospective | I: 35-37 w who attended general or high-risk prenatal visits. High risk included pre-existing chronic conditions, complications in previous pregnancies, or maternal, fetal, or placental risks in current pregnancy, or women enrolled at time of delivery without membrane rupture or hemorrhage. No exclusion criteria described. | 27 +/- 6.15 | Demographic, socio-economic, and clinical data collected through standardized questionnaires. | Recto-vaginal swabs and culture detection. | Control group might contain women with pregestational diabetes as this was not mentioned in exclusion criteria and prevalence of pregestational diabetes in GBS carriers and non-carriers was not described. | High |
| Dai et al., 2019[31] | Not reported | China | Prospective | I: Native (Chinese), 20-46 years old, 35-37 w, no sexual intercourse or antibiotic application within recent 3 months. No exclusion criteria described. | GBS+ 30.04 (3.22)<br>GBS- 30.67 (3.51) | Not reported. | Recto-vaginal swabbing followed by PCR on extracted DNA within 24 hours of collection. | Very strict inclusion criteria and did not assess prevalence of pregestational diabetes, nor differences in BMI. | High |
| Edwards et al., 2019[16] | January 1, 2003 - December 31, 2015 | USA | Retrospective cohort | All pregnant women during the timeframe were eligible. No exclusions. | GBS+ 28.0 (6.2)<br>GBS- 28.7 (6.2) | ICD codes from hospital records. | Used diagnostic codes from any time during pregnancy to determine positivity. |  | Low |
| Furfaro et al., 2019[32] | 2015-2017 | Australia | Prospective cohort | I: 16+ years old, Nulliparous/multiparous, Gestational age of less than 22 w at enrollment, understand, read, and speak English.<br>E: Highly dependent on medical care, cognitive impairment/intellectual disability, illegal drug use, antibiotic/antifungal use within 2 weeks of sample collection, multiple pregnancy (twins, etc.). | 32 (16-50) | Not reported. | Self-recto-vaginal swabbing with two swabs at each site. One swab used for multiplex PCR and the other for culture detection. |  | Low |
| Ji et al., 2019[33] | January 2016 - December 2016 | China | Population-based cohort | I: All pregnant women were screened at 35-37 w, but prior to 35 w were also tested if delivery occurred before then.<br>E: Women whose pregnancy did not result in labor. | Not provided . | Hospital records. | Recto-vaginal swab performed by physician followed by RT-PCR and culture detection. |  | Moderate |
| Manzanar es et al., 2019[34] | 2012-2014 | Spain | Case-Control | I: Delivery of a single live fetus after 26 w, BMI and GBS culture results available.<br>E: Stillbirth, or less than 26 w. | GBS+ 30.84 (5.8)<br>GBS- 30.61 (5.66) | Not reported. | Positive culture from a recto-vaginal swab at 35-37 w or GBS bacteriuria any time during pregnancy. | Urine screening replaced culture if positive any time during pregnancy, no description of culture methods. | Moderate |
| Zhu et al., 2019[35] | April 1, 2014 - March 31, 2017 | China | Retrospective cohort (population based) | I: Pregnant women 35-37 w of gestation or with preterm delivery who submitted vaginal swabs.<br>E: Women who did not undergo GBS screening, prenatal diagnosis of fetal malformation, greater or equal to three prior abortions, antibiotic usage in the week prior to admission. | GBS+ 29.7 (4.30)<br>GBS- 29.51 (4.32) | Questionnaire | Vaginal swabs followed by culture on chromogenic agar. Neonates were screened by tracheal secretion, gastric fluid, and blood sample culture. | Rectal swabs were not collected, no PCR was performed. | Moderate |
| Alfouzan et al., 2021[36] | Not reported | Lebanon | Prospective cross-sectional | Not reported. | Not reported. | Questionnaire utilized to gather sociodemographic and clinical information. | Vaginal swabs followed by culture detection. | Control group might include women with pregestational diabetes. Only vaginal swabs were collected which may underestimate GBS colonization. | Moderate |
| Huang et al., 2021[37] | June 2019-December 2020 | China | Prospective | I: No vaginal GBS colonization before pregnancy, single gestation, viable fetus, no antibiotic use during pregnancy, no sexual activity for 3 days preceding sample collection and no vaginally administered drugs or vaginal lavages 2 weeks before sample collection.<br>E: Malignant tumor, infectious diseases, comorbidities involving heart, liver lungs and other organs, genital tract malformation or incomplete medical records. | Not reported. | Determined from medical records. | Vaginal and rectal swabs followed by PCR detection. | Control group might include women with pregestational diabetes. | Moderate |
| Place et al., 2021[38] | January 2014-December 2017 | Finland | Retrospective | I: Women undergoing labor induction, singleton gestation, cephalic presentation, unfavorable cervix, intact amniotic membranes.<br>E: Women for which GBS testing was indeterminant. | 31.4 (5.4) | 2 hour 75 g oral glucose tolerance test. | Vaginal and rectal swabs PCR detection (Xpert GBS). | All women in this study had an unfavorable cervix. | Moderate |
| Del Carmen Palacios-Saucedo, et al, 2022[39] | April 2017 - December 2018 | Mexico | Prospective | Not reported. | Median and range of GBS colonized: 25 (19-37), median and range of non-colonized: 27 (14-43) | Not reported. | Recto-vaginal swabbing followed by culture and biochemical identification. | No definition of GDM. | High |
| McCoy et al., 2023[40] | December 2013-February 2017 | USA | Secondary analysis of prospective cohort study | I: singleton pregnancy and presented prior to 20 w.<br>E: Major fetal anomaly, HIV positive, history of organ transplant, chronic steroid use. | 28.6 (+/- 6.3) | Not reported. | Recto-vaginal swabs followed by culture detection per CDC guidelines | GDM diagnostic criteria and differences in severity of disease may impact findings. | Low |

### Association between gestational diabetes and maternal GBS colonization

A meta-analysis of the association between gestational diabetes and maternal rectovaginal GBS carriage revealed that women with GDM are 16% more likely to be colonized by GBS compared to women without GDM (pooled OR 1.16, CI 1.07-1.26, *P* = 0.003) (**Fig. 2**). Heterogeneity of all studies was moderate (I^2^= 34.9, *P* = 0.02). A significant driver of heterogeneity was whether the prevalence of pregestational diabetes was accounted for in the study population; sub-analysis revealed that when pregestational diabetes prevalence was not documented, and possibly present in the non-GDM control group, women with GDM had a 43% increased risk of GBS colonization (OR 1.43, CI 1.08-1.9, *P* = 0.01). Study heterogeneity was significantly greater among this subset of studies (I^2^= 67.8, *P* = 0.02). When pregestational diabetes prevalence is accounted for (thus reliably excluded from the non-GDM control group), study heterogeneity is mitigated (I^2^= 27, *P* = 0.10), and women with GDM have a 13% increased risk of GBS colonization compared to the non-diabetic control group (OR 1.13, CI 1.03-1.24, *P* = 0.01) (**Fig. 2**).

**Figure 2:**
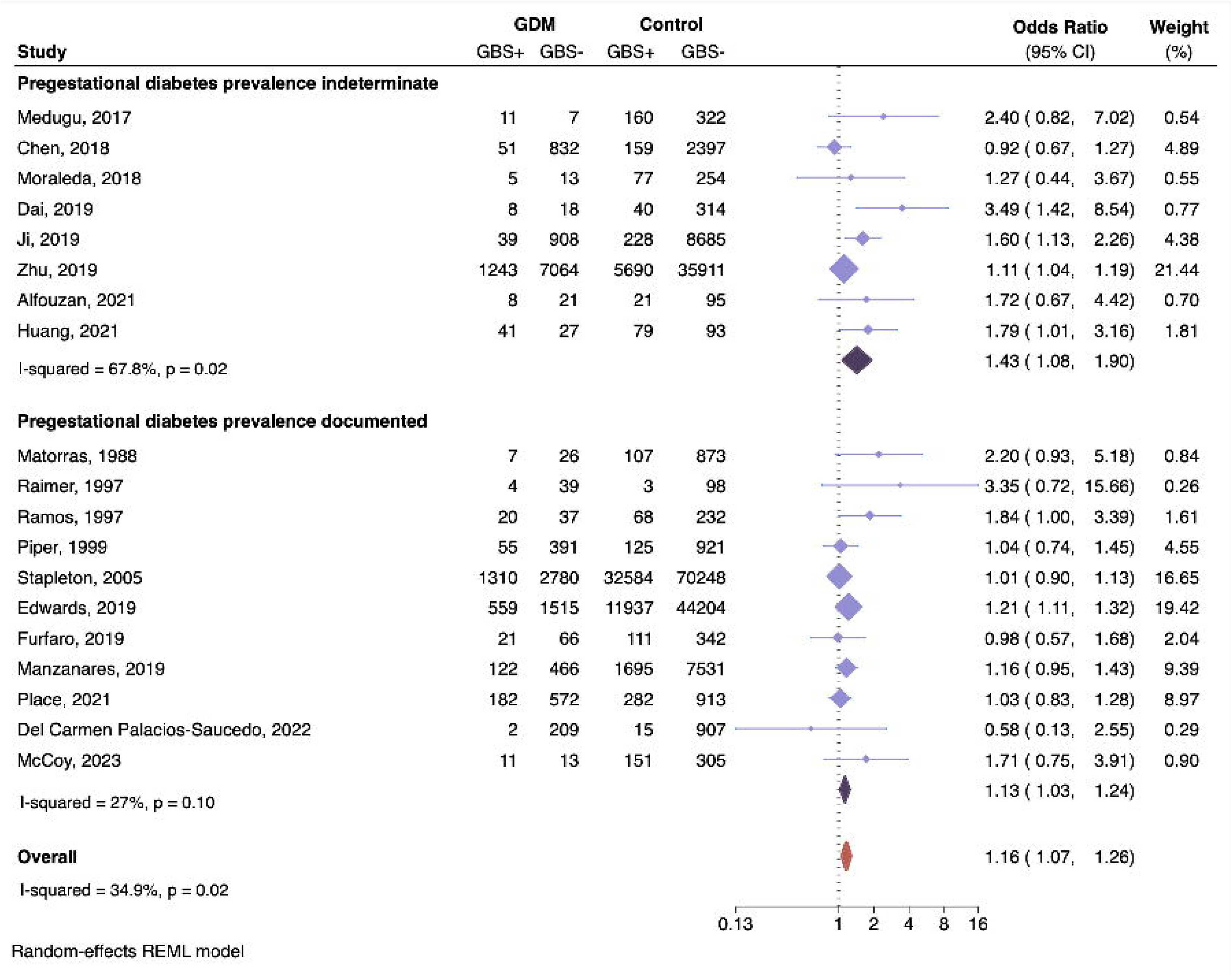
Association of gestational diabetes and GBS rectovaginal colonization. Forest plot of the association between gestational diabetes and GBS rectovaginal colonization presented as Odds Ratios (OR) for each study and respective 95% confidence intervals (CI). Studies are grouped by those that did not document (top) or did document (bottom) the prevalence of pregestational diabetes in their study population. The number of women with (GBS+) and without (GBS-) rectovaginal GBS carriage are presented for each study. The dotted black line demarcates no effect (OR = 1). The OR of individual studies are represented by light purple diamonds with shape size corresponding to the weight of the study as determined by random-effects modeling, and the paired horizontal lines indicate the 95% CI. Pooled ORs for each group are shown by the dark purple diamonds and the orange symbol represents the OR for all studies.

### Association between pregestational diabetes and maternal GBS colonization

We performed an additional sub-analysis to determine the independent association between pregestational diabetes and maternal GBS carriage, which revealed that women with pregestational diabetes have a 76% increased risk of rectovaginal GBS carriage compared to non-diabetic women (**Fig. 3**) (pooled OR 1.76, CI 1.27-2.45, *P* = 0.0008). There was a high degree of heterogeneity between studies (I^2^= 78.5%, *P* = 0.01). Appreciating distinct pathophysiology and outcomes for women with pregestational diabetes vs. gestational diabetes, we assessed differences in risk of GBS colonization. There was no significant difference in risk of GBS rectovaginal colonization based on diabetes type (**Fig. 4**); women with pregestational diabetes had a similar risk of GBS rectovaginal colonization compared to those with gestational diabetes (pooled OR 1.26, CI 0.96-1.66, *P* = 0.09). Even so, it is possible that differences will resolve with a larger sample size.

**Figure 3:**
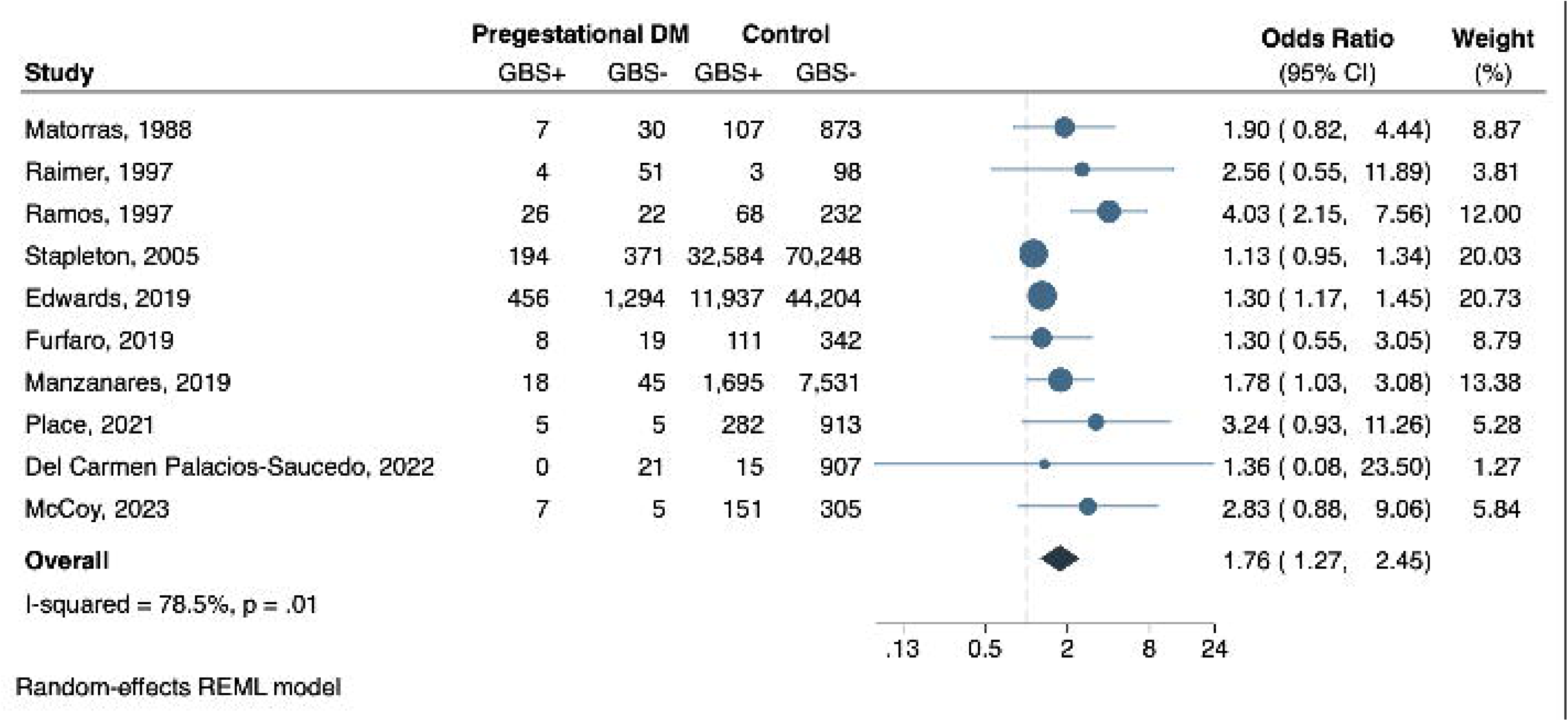
Association of pregestational diabetes and GBS rectovaginal colonization. Forest plot of the association between pregestational diabetes and GBS rectovaginal colonization presented as Odds Ratios (OR) for each study and respective 95% confidence intervals (CI). The number of women with (GBS+) and without (GBS-) rectovaginal GBS carriage are presented for each study. The dotted black line demarcates no effect (OR = 1). The OR of individual studies are represented by blue circles with shape size corresponding to the weight of the study as determined by random-effects modeling, and the paired horizontal lines indicate the 95% CI. Pooled ORs for each group are shown by the dark blue diamond.

**Figure 4:**
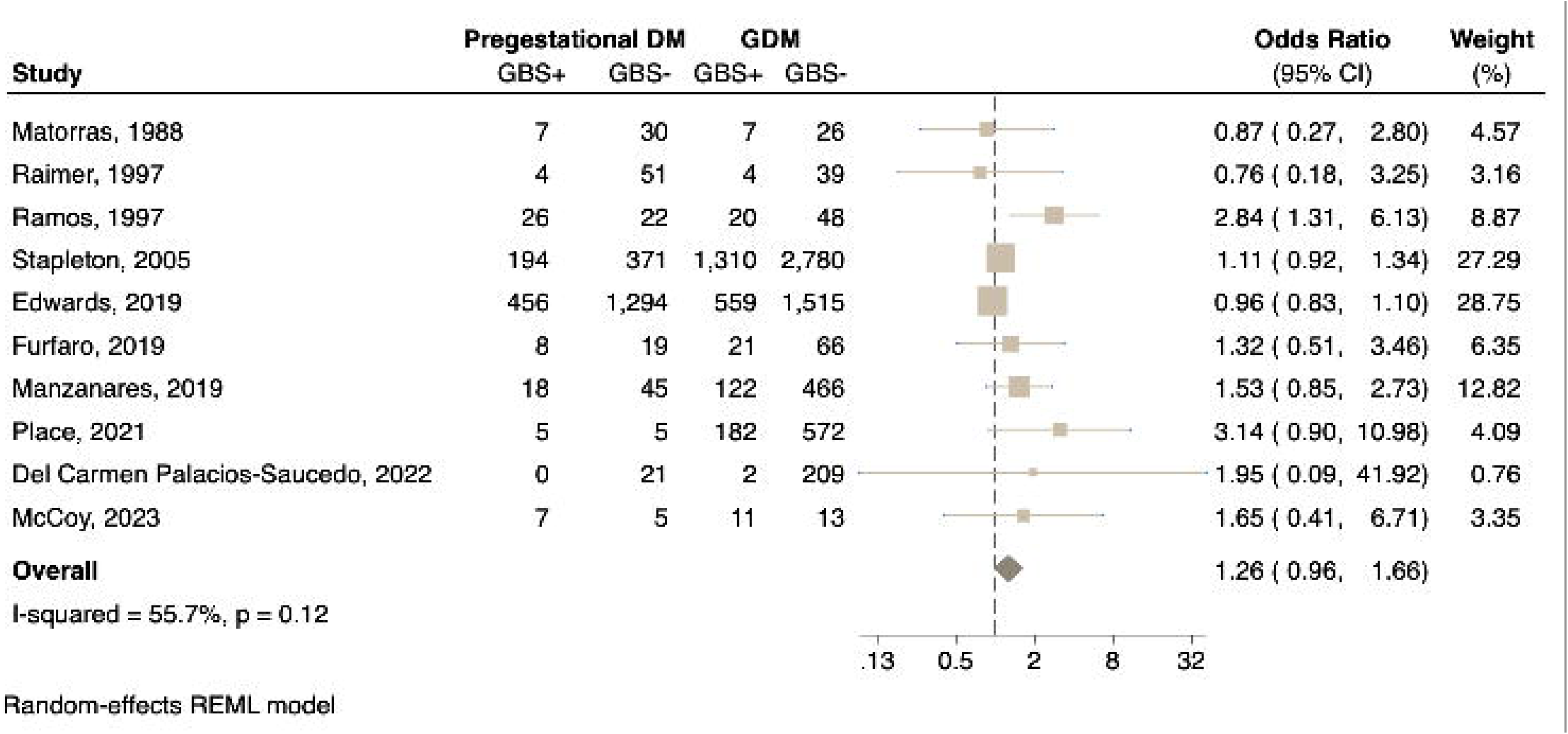
Comparison of diabetes types and associations with GBS rectobaginal colonization. Forest plot of the association between pregestational diabetes and GBS rectovaginal colonization relative to women with gestational diabetes, presented as Odds Ratios (OR) for each study and respective 95% confidence intervals (CI). The number of women with (GBS+) and without (GBS-) rectovaginal GBS carriage are presented for each study. The dotted black line demarcates no effect (OR = 1). The OR of individual studies are represented by squares with shape size corresponding to the weight of the study as determined by random-effects modeling, and the paired horizontal lines indicate the 95% CI. Pooled ORs for each group are shown by the dark beige symbol.

### Publication bias and sensitivity analysis

Visual assessment of the funnel plot (**Fig. 5**) shows asymmetrical distribution of studies, with publication bias confirmed by Egger’s test (*P* = 0.005). Sensitivity analysis included complete exclusion of studies that did not document or control for pregestational diabetic prevalence and exclusion of studies with high risk of bias. A meta-analysis was then performed on the remaining 8 studies (Raimer, Ramos, Stapleton, Edwards, Furfaro, Manzanares, Place, McCoy). Findings were robust; gestational diabetes was still associated with a 13% increased risk of rectovaginal GBS colonization (OR 1.13, 95% CI 1.02-1.25, *P* = 0.02), without significant shifts in study heterogeneity (I^2^= 35.4%, *P* = 0.08).

**Figure 5:**
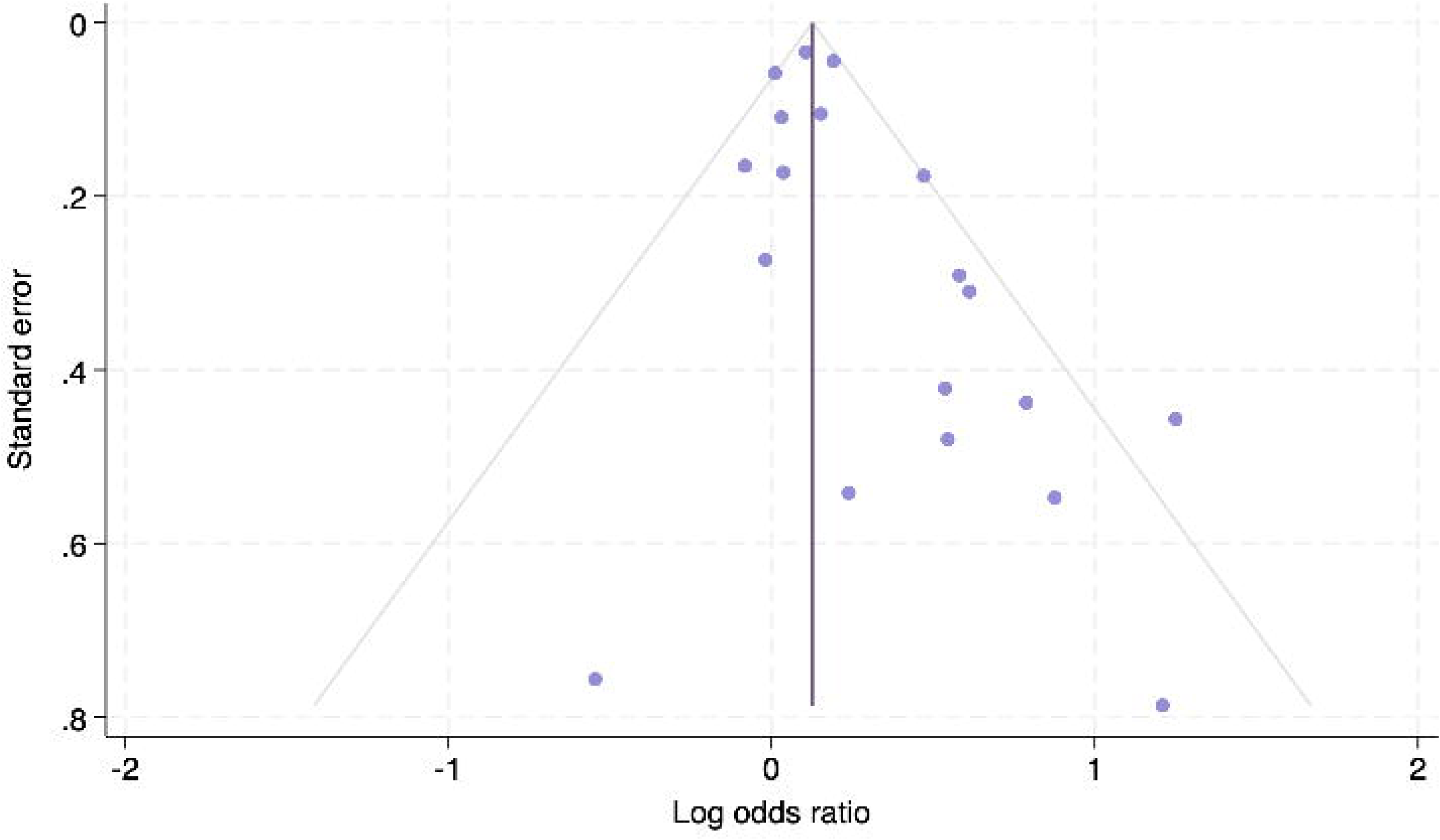
Risk of bias of included studies. Funnel plot for visual assessment of publication bias for all included studies. Circles represent individual study estimates (log odds ratio) against the respective standard error. The purple vertical line indicates the pooled OR. The gray lines mark the bounds of a pseudo 95% confidence interval.

## DISCUSSION

To our knowledge, this is the first systematic review and meta-analysis examining the association between gestational diabetes mellitus and group B streptococcal rectovaginal colonization. We also performed subgroup analyses to assess independent effects of pregestational vs. gestational diabetes on risk of maternal GBS carriage. Our meta-analysis demonstrates that women with GDM have a significant 16% greater risk of being colonized by GBS, which may in part explain the increased risk of sepsis for neonates born to mothers with GDM[13, 14]. Sub-analysis revealed that women pregestational diabetes have a 71% increased risk. There were no differences in risk based on diabetes type, which agrees with observations from prior cohort studies[17, 18].

GDM-mediated perturbations to critical host defenses such as immunity and the vaginal microbiota may mechanistically contribute to this increased susceptibility to GBS carriage. GDM leads to altered neutrophil, NK, T cell and macrophage abundance and/or activity both in the peripheral blood and at the maternal-fetal interface[10, 41–44]. While the direct role of the vaginal microbiome in propagating or limiting GBS colonization remains largely unknown, it is well-appreciated that members of the vaginal microbiota play direct and indirect roles in maintaining reproductive health and pregnancy outcomes and *Lactobacillus* dominance is considered a hallmark of an optimal vaginal community[45–49]. During pregnancy, the stability of the vaginal microbiota increases with *Lactobacillus* enrichment and overall lower alpha diversity[50, 51]. In non-diabetic pregnancy, non-*Lactobacillus* dominance or dominance by *Lactobacillus gasseri* has been associated with increased risk of GBS colonization[40, 52]. GDM disrupts the vaginal microbiota with increased diversity and enrichment of nonoptimal members including *Bacteroides*, *Klebsiella*, *Enterococcus*, and *Enterobacter* and *Staphylococcus*[11, 12, 53], of which *Staphylococcus* has been positively associated with vaginal GBS colonization[54]. Microbial communities inherited by neonates are also impacted by GDM reflected by increased colonization by *Streptococcaceae* and *Enterococcaceae* which may contribute to worse neonatal outcomes upon GBS encounter[11, 55, 56]. Indeed, in a preclinical model of GDM, we recently showed that GDM susceptibility to fetoplacental infection was driven by perturbations to maternal immunity and vaginal microbial communities in addition to pathogenic bacterial adaptations[57].

The heterogeneity observed in the 19 studies representing 10 different countries were in part explained by the presence of pregestational diabetes as a potential confounder, as revealed by sub-analyses. Other possible drivers of heterogeneity are differences in study populations including: sample size, severity of diabetes, discrepancies in access to prenatal care thereby impacting who was included in hospital or clinic-based studies, differential presence of confounding metabolic disease such as obesity, variation in inclusive representation of underrepresented or under-resourced communities, and geographical variation in GBS prevalence[1]. Differences in severity of diabetes may also explain variation between studies; a recent study found that pregnant women with better glycemic control (Hemoglobin A1c < 6.5%) had a 45% decreased risk of GBS rectovaginal colonization[58]. However, a few studies report contrasting evidence; two reports found no differences in GBS colonization status for pregnant women requiring insulin therapy[59, 60], and another study found no differences in GBS status between pregnant diabetic women requiring greater than 20 U insulin therapy vs. those requiring less than 20 U per day[61]. Thus, the association between maternal GBS colonization and diabetes severity as indicated by glycemic control and medical management (insulin treatment vs. lifestyle modification) requires further study.

Notable limitations of some of the studies incorporated in this analysis include a lack of information about GDM or pregestational diabetic severity (e.g. HbA1C or need for medical intervention), unstated GDM diagnostic criteria, potential confounding by other metabolic disorders such as obesity, and limited description of GBS detection techniques (although this was a minority of studies). However, these factors were taken into consideration in our assessment of study quality and bias risk. The presence of publication bias is another limitation and may mean that the risk is overestimated as studies with negative findings are less likely to be published. Nevertheless, sensitivity analysis revealed that the association between GDM and GBS carriage remains when high-risk and potentially confounding studies are excluded. Thus, we are confident that our findings withstand the observed limitations.

## CONCLUSIONS

Ultimately, this systematic review and meta-analysis of 19 studies representing over 260,000 women across the globe revealed gestational diabetes as a novel risk factor for maternal rectovaginal colonization by group B *Streptococcus*. Considering that up to 46% of neonates with GBS invasive disease are born to women with no currently recognized risk factors for GBS transmission, GDM may be an important risk that is not yet clinically recognized. Ultimately, expanding our knowledge of additional risk factors for GBS neonatal transmission and disease will improve strategies for screening, preventing, and treating fetal and neonatal GBS morbidity and mortality.

## Data Availability

The datasets supporting the conclusions of this article are included within the article, and in the cited studies included in the meta-analysis.

## LIST OF ABBREVIATIONS

GBS: Group B Streptococcus
IAP: Intrapartum antibiotic prophylaxis
EOGBS: Early-onset GBS disease
GDM: Gestational diabetes mellitus

## DECLARATIONS

### Authors’ contributions

KAP and VME conceived and designed the study. VME, JJZ, and ZH, performed systematic review and analyses. VME and KAP analyzed data and interpreted results. KAP secured funding. VME and KAP drafted the manuscript. All authors reviewed and edited the manuscript.

## REFERENCES

1. Gonçalves BP, Procter SR, Paul P, Chandna J, Lewin A, Seedat F, Koukounari A, Dangor Z, Leahy S, Santhanam S et al: Group B streptococcus infection during pregnancy and infancy: estimates of regional and global burden. Lancet Glob Health 2022, 10(6):e807–e819.

2. Kwatra G, Cunnington MC, Merrall E, Adrian PV, Ip M, Klugman KP, Tam WH, Madhi SA: Prevalence of maternal colonisation with group B streptococcus: a systematic review and meta-analysis. Lancet Infect Dis 2016, 16(9):1076–1084.

3. Russell NJ, Seale AC, O’Driscoll M, O’Sullivan C, Bianchi-Jassir F, Gonzalez-Guarin J, Lawn JE, Baker CJ, Bartlett L, Cutland C et al: Maternal Colonization With Group B Streptococcus and Serotype Distribution Worldwide: Systematic Review and Meta-analyses. Clin Infect Dis 2017, 65(suppl_2):S100–s111.

4. Schrag SJ, Farley MM, Petit S, Reingold A, Weston EJ, Pondo T, Hudson Jain J, Lynfield R: Epidemiology of Invasive Early-Onset Neonatal Sepsis, 2005 to 2014. Pediatrics 2016, 138(6).

5. Hasperhoven GF, Al-Nasiry S, Bekker V, Villamor E, Kramer B: Universal screening versus risk-based protocols for antibiotic prophylaxis during childbirth to prevent early-onset group B streptococcal disease: a systematic review and meta-analysis. Bjog 2020, 127(6):680–691.

6. Heath PT, Balfour GF, Tighe H, Verlander NQ, Lamagni TL, Efstratiou A: Group B streptococcal disease in infants: a case control study. Arch Dis Child 2009, 94(9):674–680.

7. Trijbels-Smeulders M, de Jonge GA, Pasker-de Jong PC, Gerards LJ, Adriaanse AH, van Lingen RA, Kollée LA: Epidemiology of neonatal group B streptococcal disease in the Netherlands before and after introduction of guidelines for prevention. Arch Dis Child Fetal Neonatal Ed 2007, 92(4):F271–276.

8. Wang H, Li N, Chivese T, Werfalli M, Sun H, Yuen L, Hoegfeldt CA, Elise Powe C, Immanuel J, Karuranga S et al: IDF Diabetes Atlas: Estimation of Global and Regional Gestational Diabetes Mellitus Prevalence for 2021 by International Association of Diabetes in Pregnancy Study Group’s Criteria. Diabetes Res Clin Pract 2022, 183:109050.

9. McIntyre HD, Catalano P, Zhang C, Desoye G, Mathiesen ER, Damm P: Gestational diabetes mellitus. Nat Rev Dis Primers 2019, 5(1):47.

10. McElwain CJ, McCarthy FP, McCarthy CM: Gestational Diabetes Mellitus and Maternal Immune Dysregulation: What We Know So Far. Int J Mol Sci 2021, 22(8).

11. Wang J, Zheng J, Shi W, Du N, Xu X, Zhang Y, Ji P, Zhang F, Jia Z, Wang Y et al: Dysbiosis of maternal and neonatal microbiota associated with gestational diabetes mellitus. Gut 2018, 67(9):1614–1625.

12. Cortez RV, Taddei CR, Sparvoli LG, Ângelo AGS, Padilha M, Mattar R, Daher S: Microbiome and its relation to gestational diabetes. Endocrine 2019, 64(2):254–264.

13. Håkansson S, Källén K: Impact and risk factors for early-onset group B streptococcal morbidity: analysis of a national, population-based cohort in Sweden 1997-2001. Bjog 2006, 113(12):1452–1458.

14. Medugu N, Iregbu KC, Parker RE, Plemmons J, Singh P, Audu LI, Efetie E, Davies HD, Manning SD: Group B streptococcal colonization and transmission dynamics in pregnant women and their newborns in Nigeria: implications for prevention strategies. Clinical Microbiology and Infection 2017, 23(9):673.e679–673.e616.

15. Piper JM, Georgiou S, Xenakis EM, Langer O: Group B streptococcus infection rate unchanged by gestational diabetes. Obstet Gynecol 1999, 93(2):292–296.

16. Edwards JM, Watson N, Focht C, Wynn C, Todd CA, Walter EB, Heine RP, Swamy GK: Group B Streptococcus (GBS) Colonization and Disease among Pregnant Women: A Historical Cohort Study. Infect Dis Obstet Gynecol 2019, 2019:5430493.

17. Matorras R, Garcia-Perea A, Usandizaga JA, Omenaca F: Recto-vaginal colonization and urinary tract infection by group B streptococcus in the pregnant diabetic patient. Acta Obstetricia et Gynecologica Scandinavica 1988, 67(7):617–620.

18. Ramos E, Gaudier FL, Hearing LR, Del Valle GO, Jenkins S, Briones D: Group B streptococcus colonization in pregnant diabetic women. Obstet Gynecol 1997, 89(2):257–260.

19. Bey M, Pastorek Ii JG, Miller Jr JM: Group B streptococcal colonization in the diabetic gravida patient. American Journal of Perinatology 1992, 9(5-6):425–427.

20. Chen X, Cao S, Fu X, Ni Y, Huang B, Wu J, Chen L, Huang S, Cao J, Yu W, Ye H: The risk factors for Group B Streptococcus colonization during pregnancy and influences of intrapartum antibiotic prophylaxis on maternal and neonatal outcomes. BMC Pregnancy Childbirth 2023, 23(1):207.

21. Farideh A, Abdolkarim H, Naderi Nasab M: Comparison of Group B Streptococcal Colonization in the Pregnant Diabetic and Non-Diabetic Women. Acta Medica Iranica 2007, 47(2).

22. Siqueira F, Ferreira EM, de Matos Calderon I, Dias A: Prevalence of colonisation by group B streptococcus in pregnant patients in Taguatinga, Federal District, Brazil: a cross-sectional study. Arch Gynecol Obstet 2019, 299(3):703–711.

23. Lukic A, Napoli A, Santino I, Bianchi P, Nobili F, Ciampittiello G, Nardone MR, Santomauro M, Di Properzio M, Caserta D: Cervicovaginal bacteria and fungi in pregnant diabetic and non-diabetic women: a multicenter observational cohort study. Eur Rev Med Pharmacol Sci 2017, 21(10):2303–2315.

24. Higgins JP, Altman DG, Gøtzsche PC, Jüni P, Moher D, Oxman AD, Savovic J, Schulz KF, Weeks L, Sterne JA: The Cochrane Collaboration’s tool for assessing risk of bias in randomised trials. Bmj 2011, 343:d5928.

25. Higgins JP, Thompson SG: Quantifying heterogeneity in a meta-analysis. Stat Med 2002, 21(11):1539–1558.

26. 2022-2023 World Bank Country Lending Group guidelines [https://worldpopulationreview.com/country-rankings/middle-income-countries]

27. Raimer K, O’Sullivan MJ: Influence of diabetes on group B Streptococcus colonization in the pregnant patient. J Matern Fetal Med 1997, 6(2):120–123.

28. Stapleton RD, Kahn JM, Evans LE, Critchlow CW, Gardella CM: Risk factors for group B streptococcal genitourinary tract colonization in pregnant women. Obstet Gynecol 2005, 106(6):1246–1252.

29. Chen J, Fu J, Du W, Liu X, Rongkavilit C, Huang X, Wu Y, Wang Y, McGrath E: Group B streptococcal colonization in mothers and infants in western China: Prevalences and risk factors. BMC Infectious Diseases 2018, 18(1).

30. Moraleda C, Benmessaoud R, Esteban J, López Y, Alami H, Barkat A, Houssain T, Kabiri M, Bezad R, Chaacho S et al: Prevalence, antimicrobial resistance and serotype distribution of group B streptococcus isolated among pregnant women and newborns in Rabat, Morocco. Journal of Medical Microbiology 2018, 67(5):652–661.

31. Dai W, Zhang Y, Xu Y, Zhu M, Rong X, Zhong Q: The effect of group B streptococcus on maternal and infants’ prognosis in Guizhou, China. Bioscience Reports 2019, 39(12).

32. Furfaro LL, Nathan EA, Chang BJ, Payne MS: Group B streptococcus prevalence, serotype distribution and colonization dynamics in Western Australian pregnant women. Journal of Medical Microbiology 2019, 68(5):728–740.

33. Ji Y, Zhao C, Ma XX, Peppelenbosch MP, Ma Z, Pan Q: Outcome of a screening program for the prevention of neonatal early-onset group B Streptococcus infection: A population-based cohort study in inner Mongolia, China. Journal of Medical Microbiology 2019, 68(5):803–811.

34. Manzanares S, Zamorano M, Naveiro-Fuentes M, Pineda A, Rodríguez-Granger J, Puertas A: Maternal obesity and the risk of group B streptococcal colonisation in pregnant women. Journal of Obstetrics and Gynaecology 2019, 39(5):628–632.

35. Zhu Y, Huang J, Lin XZ, Chen C: Group B Streptococcus Colonization in Late Pregnancy and Invasive Infection in Neonates in China: A Population-Based 3-Year Study. Neonatology 2019, 115(4):301–309.

36. Alfouzan W, Gaddar N, Dhar R, Rabaan AA: A study of group B streptococcus in pregnant women in Lebanon: Prevalence, risk factors, vaginal flora and antimicrobial susceptibility. Infezioni in Medicina 2021, 29(1):85–93.

37. Huang J, Zheng L, Su Y, Wang F, Kong H, Chang Y, Xin H: Effects of group B streptococcus infection on vaginal micro-ecology and pregnancy outcomes of pregnant women in late pregnancy. European Journal of Obstetrics and Gynecology and Reproductive Biology 2021, 267:274–279.

38. Place K, Rahkonen L, Nupponen I, Kruit H: Vaginal streptococcus B colonization is not associated with increased infectious morbidity in labor induction. Acta Obstetricia et Gynecologica Scandinavica 2021, 100(8):1501–1510.

39. Del Carmen Palacios-Saucedo G, Rivera-Morales LG, Vazquez-Guillen JM, Caballero-Trejo A, Mellado-Garcia MC, Flores-Flores AS, Gonzalez-Navarro JA, Herrera-Rivera CG, Osuna-Rosales LE, Hernandez-Gonzalez JA et al: Genomic analysis of virulence factors and antimicrobial resistance of group B Streptococcus isolated from pregnant women in northeastern Mexico. PLoS ONE 2022, 17(3 March).

40. McCoy JA, Burris HH, Gerson KD, McCarthy C, Ravel J, Elovitz MA: Cervicovaginal Microbial-Immune State and Group B Streptococcus Colonization in Pregnancy. Am J Perinatol 2023.

41. Stoikou M, Grimolizzi F, Giaglis S, Schäfer G, van Breda SV, Hoesli IM, Lapaire O, Huhn EA, Hasler P, Rossi SW, Hahn S: Gestational Diabetes Mellitus Is Associated with Altered Neutrophil Activity. Front Immunol 2017, 8:702.

42. Hara Cde C, França EL, Fagundes DL, de Queiroz AA, Rudge MV, Honorio-França AC, Calderon Ide M: Characterization of Natural Killer Cells and Cytokines in Maternal Placenta and Fetus of Diabetic Mothers. J Immunol Res 2016, 2016:7154524.

43. Mihalic Z, Kindler O, Raftopoulou S, Santiso A, Wadsack C, Heinemann A, Kargl J: Gestational diabetes mellitus dysregulates the PD-1/PD-L1 axis at the feto-maternal interface. bioRxiv 2023:2023.2001.2025.525478.

44. Kang YE, Yi HS, Yeo MK, Kim JT, Park D, Jung Y, Kim OS, Lee SE, Kim JM, Joung KH et al: Increased Pro-Inflammatory T Cells, Senescent T Cells, and Immune-Check Point Molecules in the Placentas of Patients With Gestational Diabetes Mellitus. J Korean Med Sci 2022, 37(48):e338.

45. Greenbaum S, Greenbaum G, Moran-Gilad J, Weintraub AY: Ecological dynamics of the vaginal microbiome in relation to health and disease. Am J Obstet Gynecol 2019, 220(4):324–335.

46. Dunlop AL, Satten GA, Hu YJ, Knight AK, Hill CC, Wright ML, Smith AK, Read TD, Pearce BD, Corwin EJ: Vaginal Microbiome Composition in Early Pregnancy and Risk of Spontaneous Preterm and Early Term Birth Among African American Women. Front Cell Infect Microbiol 2021, 11:641005.

47. Tabatabaei N, Eren AM, Barreiro LB, Yotova V, Dumaine A, Allard C, Fraser WD: Vaginal microbiome in early pregnancy and subsequent risk of spontaneous preterm birth: a case-control study. Bjog 2019, 126(3):349–358.

48. Ravel J, Gajer P, Abdo Z, Schneider GM, Koenig SS, McCulle SL, Karlebach S, Gorle R, Russell J, Tacket CO et al: Vaginal microbiome of reproductive-age women. Proc Natl Acad Sci U S A 2011, 108 Suppl 1(Suppl 1):4680–4687.

49. Zhou X, Bent SJ, Schneider MG, Davis CC, Islam MR, Forney LJ: Characterization of vaginal microbial communities in adult healthy women using cultivation-independent methods. Microbiology (Reading) 2004, 150(Pt 8):2565–2573.

50. Freitas AC, Chaban B, Bocking A, Rocco M, Yang S, Hill JE, Money DM: The vaginal microbiome of pregnant women is less rich and diverse, with lower prevalence of Mollicutes, compared to non-pregnant women. Sci Rep 2017, 7(1):9212.

51. DiGiulio DB, Callahan BJ, McMurdie PJ, Costello EK, Lyell DJ, Robaczewska A, Sun CL, Goltsman DS, Wong RJ, Shaw G et al: Temporal and spatial variation of the human microbiota during pregnancy. Proc Natl Acad Sci U S A 2015, 112(35):11060–11065.

52. Rosen GH, Randis TM, Desai PV, Sapra KJ, Ma B, Gajer P, Humphrys MS, Ravel J, Gelber SE, Ratner AJ: Group B Streptococcus and the Vaginal Microbiota. J Infect Dis 2017, 216(6):744–751.

53. Rafat D, Singh S, Nawab T, Khan F, Khan AU, Khalid S: Association of vaginal dysbiosis and gestational diabetes mellitus with adverse perinatal outcomes. Int J Gynaecol Obstet 2022, 158(1):70–78.

54. Shabayek S, Abdellah AM, Salah M, Ramadan M, Fahmy N: Alterations of the vaginal microbiome in healthy pregnant women positive for group B Streptococcus colonization during the third trimester. BMC Microbiol 2022, 22(1):313.

55. He Z, Wu J, Xiao B, Xiao S, Li H, Wu K: The Initial Oral Microbiota of Neonates Among Subjects With Gestational Diabetes Mellitus. Front Pediatr 2019, 7:513.

56. Chen T, Qin Y, Chen M, Zhang Y, Wang X, Dong T, Chen G, Sun X, Lu T, White RA, 3rd et al: Gestational diabetes mellitus is associated with the neonatal gut microbiota and metabolome. BMC Med 2021, 19(1):120.

57. Mercado-Evans V, Mejia ME, Zulk JJ, Ottinger S, Hameed Z, Serchejian C, Marunde MG, Robertson CM, Ballard MB, Korotkova N et al: Gestational diabetes augments group B <EM>Streptococcu</EM>s perinatal infection through disruptions in maternal immunity and the vaginal microbiota. bioRxiv 2023:2023.2006.2023.546252.

58. Field C, Bank TC, Spees CK, Germann K, Landon MB, Gabbe S, Grobman WA, Costantine MM, Venkatesh KK: Association between glycemic control and group B streptococcus colonization among pregnant individuals with pregestational diabetes. Am J Reprod Immunol 2023, 90(4):e13779.

59. Ramos E, Gaudier FL, Hearing LR, Del Valle GO, Jenkins S, Briones D: Group B streptococcus colonization in pregnant diabetic women. Obstetrics and Gynecology 1997, 89(2):257–260.

60. Pykało-Gawińska D, Zaręba-Szczudlik J, Gawiński C, Stępień A, Dobrowolska-Redo A, Malinowska-Polubiec A, Romejko-Wolniewicz E: Gestational weight gain and glycemic control in GDM patients with positive genital culture. Taiwanese Journal of Obstetrics and Gynecology 2021, 60(2):262–265.

61. Bey M, Pastorek JG, 2nd, Miller JM, Jr.: Group B streptococcal colonization in the diabetic gravida patient. Am J Perinatol 1992, 9(5-6):425–427.

