## Supplemental Material for "Gestational diabetes as a risk factor for GBS maternal rectovaginal colonization: a systematic review and meta-analysis"

**SUPPLEMENTAL MATERIALS**

**Supplemental table 1: Studies excluded upon full-text appraisal.**

| Rationale for study exclusion | Studies excluded |
|---|---|
| Did not distinguish diabetes type *n* = 22 | Arias et al., 2022, Akhlaghi et al., 2007, Bey et al., 1992, Chen et al., 2019, Chen et al., 2023, Embil et al. 1978, Ernest et al., 2015, Gopal Rao et al., 2019, Herrera et al., 2023, Horvath et al., 2013, Horvath et al., 2013, Kessous et al., 2012, Lukic et al., 2017, Moghaddam et al., 2010, Mulla et al., 2012, Musleh et al., 2018, Najmi et al., 2013, Obata-Yasuoka, 2012, Rick et al., 2017, Shah et al., 2011, Strus et al., 2009, Zhang, 2023 |
| Did not assess GDM as a risk factor *n* = 18 | Akman et al., 2001, Alzuheiri et al., 2021, Belhadi et al., 2020, Berikopoulou et al., 2021, Botelho et al., 2018, Chaudhary et al., 2015, Dadi et al., 2022, Dahl et al., 2003, Elvedi-Gašparović et al., 2008, Ji et al., 2017, Jones et al., 2006, Khamching et al., 2017, Khan et al., 2015, Nahaei et al., 2007, Philipson et al. 1996, Pykało-Gawińska et al., 2021, Tor-udom et al., 2006, Werawatakul et al., 2001 |
| Incomplete information *n* = 3 | Clouse et al., 2019, Regan et al., 1991, Siqueira et al., 2019 |
| Exclusion and inclusion criteria not reported *n* = 1 | Hammoud et al., 2002 |
| Article not in English *n* = 1 | Jerbi et al., 2007 |
| Article not accessible online *n* = 1 | Prośniewska, M. et al., 2014 |

**Supplemental table 2: Risk of bias and quality assessment scale.**

| | |
|---|---|
| **SELECTION BIAS** | |
| **Representativeness of the exposed group** | Score & Comments |
| 0- Representative of the general population | |
| 1- Moderately limited in its representation of the general population | |
| 2- Only representative of a select population | |
| 3- No description of the sampling strategy (inclusion/exclusion criteria) | |
| **Representativeness of the control group** | Score & Comments |
| 0- Representative of the general population | |
| 1- Moderately limited in its representation of the general population | |
| 2- Only representative of a select population | |
| 3- No description of the sampling strategy (inclusion/exclusion criteria) | |
| **Sample size** | Score & Comments |
| 0- Satisfactory or justified: power calculations reported or $n \geq 488$ | |
| 1- Not satisfactory or justified | |
| **COMPARABILITY** | |
| **Comparability of the cohorts** | Score & Comments |
| 0- Exposed and non-exposed matched in design or confounders adjusted for in analysis | |
| 1- Cohorts are not matched and confounders not adjusted for in analyses, or information not provided | |
| **MEASUREMENT BIAS** | |
| **Ascertainment of group B Streptococcal carriage** | Score & Comments |
| 0- Sampling, culturing and identification thoroughly performed and described with substantial adherence to CDC guidelines | |
| 1- Sampling, culturing and identification thoroughly performed and described but without substantial adherence to CDC guidelines | |
| 2- Limited or no description | |
| **Ascertainment of GDM exposure** | Score & Comments |
| 0- Diagnosis reported from medical professional or medical records with diagnostic criteria specified | |
| 1- Diagnosis reported from medical professional or medical records with diagnostic criteria not specified | |
| 2- Self-reporting/questionnaires | |
| 3- No description | |
| **POTENTIAL OF BIAS FROM OTHER METABOLIC DISEASES** | |
| **Presence of other metabolic disease(s) in cohort** | Score & Comments |

| |
|---|
| 0- Prevalence of obesity and pregestational diabetes in exposed and nonexposed groups is provided, or other metabolic diseases were part of exclusion criteria |
| 1- Prevalence of obesity and pregestational diabetes not provided or not part of exclusion criteria |

**Supplemental table 3: Quality and risk of bias scores for each study.**

| Study | Selection bias | Cohort Comparability | Measurement bias | Bias from other metabolic diseases | Total Score | Overall Bias |
|---|---|---|---|---|---|---|
| Matorras, 1988 | 6 | 1 | 0.5 | 1 | 8.5 | High |
| Raimer, 1997 | 2.5 | 1 | 1 | 1 | 5.5 | Mod.[a] |
| Ramos, 1997 | 2 | 0 | 1 | 0 | 3 | Low |
| Piper et al., 1999 | 4 | 1 | 0.5 | 1 | 6.5 | High |
| Stapleton et al., 2005 | 0 | 0 | 2.5 | 0 | 2.5 | Low |
| Medugu et al., 2017 | 0 | 1 | 1.5 | 1 | 3.5 | Mod. |
| Chen et al., 2018 | 0 | 0 | 1 | 1 | 2 | Low |
| Moraleda et al., 2018 | 1 | 1 | 3.5 | 1 | 6.5 | High |
| Dai et al., 2019 | 3 | 1 | 2 | 1 | 7 | High |
| Edwards et al., 2019 | 0 | 0 | 2 | 1 | 3 | Low |
| Furfaro et al., 2019 | 0 | 0 | 1 | 0 | 1 | Low |
| Ji et al., 2019 | 2 | 1 | 1.5 | 1 | 5.5 | Mod. |
| Manzanares et al., 2019 | 2 | 0 | 3 | 0 | 5 | Mod. |
| Zhu et al., 2019 | 0 | 0 | 3.5 | 1 | 4.5 | Mod. |
| Alfouzan et al., 2021 | 3 | 0 | 2 | 1 | 6 | Mod. |
| Huang et al., 2021 | 3 | 0 | 2 | 1 | 6 | Mod. |
| Place et al., 2021 | 4 | 0 | 0.5 | 1 | 5.5 | Mod. |
| Del Carmen Palacios-Saucedo, et al, 2022 | 4 | 1 | 3 | 0.5 | 8.5 | High |

| | | | | | | |
|---|---|---|---|---|---|---|
| McCoy et al., 2023 | 2 | 0 | 1 | 0 | 3 | Low |
| Total possible points for category | *7* | *1* | *5* | *1* | *14* | - |

[a]Mod. = moderate
